# Screening and Diagnosis Trends for Primary Aldosteronism: A Longitudinal Nationwide Cohort Study of 7.8 Million People

**DOI:** 10.1101/2025.11.13.25340212

**Authors:** Cheng-Hsuan Tsai, Yu-Ching Chang, Zheng-Wei Chen, Stefanie Parisien-La Salle, Jenifer M Brown, Anand Vaidya, Vin-Cent Wu, Yen-Hung Lin

## Abstract

**Background:** Primary aldosteronism (PA) is a common, treatable cause of hypertension for which screening is widely recommended but rarely performed in clinical practice. However, real-world screening and diagnosis patterns across the entire hypertensive population remain unknown. This study aimed to delineate the 22-year state of nationwide PA screening and diagnosis rates among all hypertensive population in Taiwan

**Methods:** In this nationwide retrospective cohort study from 2001 to 2022, we identified all patients with hypertension using a national health insurance database. We calculated annual PA screening and diagnosis rates, with particular focus on high-risk subgroups, including patients with resistant hypertension, early-onset hypertension, hypokalemia, and other comorbidities warranting screening.

**Results:** Among 7.8 million patients with hypertension, a total of 4.4% received PA screening during the study period. The annual PA screening rate increased from 0.26% in 2001 to 0.75% in 2022 (*p* < 0.001) yet remained markedly low. In 2022, only 1.0% of patients with resistant hypertension, 3.0% with early-onset hypertension, and 3.6% with hypokalemia underwent screening. The diagnostic yield of PA showed a slight decrease over time, fluctuating between 8.0% and 6.7% (*p* = 0.006).

**Conclusions:** Despite an increase in PA screening over the past two decades, absolute rates remain critically low, falling far short of guideline recommendations, especially in high-risk groups. Our findings quantify a major implementation gap between evidence and clinical practice. As international guidelines are shifting towards broader and simpler screening protocols, there is an urgent need to improve the detection of this common and actionable condition.

**Perspective:** *What is known?:*

- Primary aldosteronism (PA) is a common, clinically important, and treatable cause of hypertension. Its adverse cardiovascular effects arise from renin independent aldosterone excess and chronic mineralocorticoid receptor overactivation.
- Current guidelines historically recommended screening only in high-risk groups, but recent expert consensus and international guidelines increasingly support broader and even universal screening among all hypertensive patients.
- Prior epidemiologic studies have examined PA screening almost exclusively within selected high-risk subgroups. No study has comprehensively evaluated real-world screening patterns across an entire hypertensive population.

*What is new?:*

- Using a 22-year nationwide cohort of nearly 8 million hypertensive patients, this is the first study to comprehensively quantify real-world trends in PA screening and diagnosis in Taiwan.
- Annual screening rates nearly tripled but remained profoundly low (<4%), even in guideline-recommended high-risk groups such as resistant hypertension, early-onset hypertension, and hypokalemia.
- Hypertensive patients receiving care at tertiary medical centers or in highly urbanized areas were more likely to undergo screening.

*What is next?:*

- The persistent under-screening of PA represents a major implementation gap, resulting in missed opportunities for targeted treatment and long-term cardiovascular risk reduction.
- Closing this gap will require a paradigm shift that includes broader screening strategies, decentralizing testing to primary care, and adopting simplified diagnostic pathways aligned with contemporary guidelines.
- Future efforts should evaluate the impact of these simplified approaches on diagnosis rates, treatment implementation, and cardiovascular outcomes, particularly as global practice moves toward earlier and more inclusive detection of renin-independent aldosteronism.

## Introduction

Primary aldosteronism (PA) is the most common and treatable cause of secondary hypertension(1–3). Its prevalence is substantial, estimated to affect up to 25% of the general hypertensive population(4–10). The clinical importance of distinguishing PA from essential hypertension lies in its distinct pathophysiology which is characterized by renin-independent aldosterone production(11,12). This dysregulated aldosterone production, coupled with mineralocorticoid receptor overactivation, contributes not only to elevated blood pressure but also to a significantly higher cardiometabolic risk compared to essential hypertension(3,13–15). Importantly, aldosterone-targeted therapies can markedly reduce these risks(16,17). However, identifying and screening affected individuals remain challenging, leading to widespread underdiagnosis.

In recognition of PA’s prevalence and associated risks, clinical practice guidelines from major societies have long recommended targeted screening in high-risk patients, including those with resistant hypertension, early-onset hypertension, and hypertension with hypokalemia(18–20). More recently, the understanding of PA has evolved from a discrete disease to a broad spectrum of renin-independent aldosterone production. This paradigm shift has led some experts and guidelines to advocate for universal screening for all patients with hypertension(20–22). However, whether current real-world practice aligns with this universal approach remains unknown. Previous studies have focused almost exclusively on screening rates within high-risk patients(23–25); none have comprehensively evaluated the state of screening across the nationwide hypertensive population.

Therefore, this study leveraged the nationwide health insurance database from Taiwan to delineate two decades of PA screening and diagnostic trends, quantify temporal changes, and critically assess real-world screening practices with a focus on high-risk subgroups. We therefore aimed to deliver a comprehensive, long-term nationwide analysis that offers valuable insights into the challenges of implementing PA screening for all hypertensive patients in clinical practice.

## Methods

### Data source

This retrospective, nationwide, population-based cohort study was conducted using data from the Taiwan National Health Insurance Research Database (NHIRD). The National Health Insurance (NHI) program in Taiwan is a single-payer system with mandatory enrollment which was implemented in 1995, now covering over 99.9% of the nation’s population. The NHIRD contains comprehensive, anonymized data on all beneficiaries, including diagnoses, procedures, and prescription drug claims. Access to the NHIRD has now shifted to the Health and Welfare Data Science Center of the Ministry of Health and Welfare, enabling full-population analyses and linkage with other databases (e.g., the National Death Registry) to expand research potential. Further details of the NHIRD can be found in prior reports(26–28). As all personal information in the NHIRD is de-identified, this study was granted a waiver of full review by the Institutional Review Board of National Taiwan University Hospital, and the need for informed consent was waived (202508048W).

### Study overview

The study was structured in three sequential steps. First, we quantified the overall screening proportion, defined as the proportion of hypertensive patients who underwent PA screening at any time during the study period. Second, we evaluated annual screening rates, calculated as the number of patients screened in each calendar year divided by the total number of living hypertensive patients in that year, both in the overall population and within predefined high-risk groups. We also examined patient- and system-level factors associated with the likelihood of screening. Third, we assessed the diagnostic yield of PA, defined annually as the proportion of screened patients who received a confirmed diagnosis according to contemporaneous clinical PA guidelines, again evaluated in the overall population and in high-risk groups.

### Study population

For this study, we obtained approved access to NHIRD data for the period 2000–2023. In Taiwan, the International Classification of Diseases, Ninth Revision, Clinical Modification (ICD-9-CM) was used for diagnostic coding until December 31, 2015, and both ICD-9-CM and the Tenth Revision, Clinical Modification (ICD-10-CM) have been used thereafter. All patients with a new-onset diagnosis of hypertension were initially identified between January 1, 2001, and December 31, 2022. Patients with hypertension diagnosed in 2000 were not included to restrict the cohort to newly diagnosed cases. Previous validation studies in the NHIRD have confirmed the accuracy of algorithms used to define hypertension, with an accuracy of 93% for the definition based solely on diagnoses from at least two outpatient visits or one hospital admission, and 95% when based solely on antihypertensive medication use(29). To enhance diagnostic accuracy, we defined hypertension as the presence of a diagnosis plus the use of any class of antihypertensive medication for at least three months. Patients with missing information on date of birth, sex, monthly income, zip-code information including residence or institution of hypertension diagnosed were excluded from the analysis.

### Definition of PA Screening and Diagnosis

Screening for PA was identified by claims for both a plasma renin (either plasma renin activity or direct renin concentration) and a plasma aldosterone concentration test conducted on the same day. PA screening was ascertained through Taiwan NHI reimbursement codes. A confirmed diagnosis of PA was defined operationally by the presence of a diagnostic code for PA (at least two outpatient visits or one hospital admission) in conjunction with either a subsequent claim for adrenalectomy or a persistent prescription for a mineralocorticoid receptor antagonist (MRA), such as spironolactone, for at least three consecutive months within 730 days after the PA screening(30). While this operational definition may not capture all diagnosed cases (e.g., those not initiated on targeted therapy), it may enhance the specificity of our cohort of ‘confirmed and treated PA’, thereby providing a conservative but robust estimate of diagnostic trends

The potentially high-risk subgroups in this study included patients with early-onset hypertension (defined as hypertension diagnosed before the age of 40), hypertension with hypokalemia, resistant hypertension, chronic kidney disease, obstructive sleep apnea, ischemic stroke, and intracranial hemorrhage. In this study, resistant hypertension was operationalized in two complementary ways. For the baseline characteristics, resistant hypertension at baseline was defined as the use of four or more antihypertensive agents within three months after the initial diagnosis of hypertension. For the analysis of annual screening rates and diagnostic yields, resistant hypertension in the year was defined according to the cumulative exposure to antihypertensive therapy, calculated as the total number of prescription days for medications from the eight major antihypertensive drug classes, divided by 270. A value equal to or greater than four was considered resistant hypertension. This annualized definition allowed patients to newly enter or leave the resistant hypertension subgroup over time, reflecting changes in treatment intensity. Hypokalemia was defined as having at least two outpatient diagnoses or one inpatient diagnosis at any time prior to the index date; chronic kidney disease and obstructive sleep apnea were defined as at least two outpatient diagnoses or one inpatient diagnosis within the past year; and stroke was defined as any prior hospitalization with the corresponding diagnosis. We also extracted information on other covariates, including personal monthly income (classified into tertiles), urbanization level of residence, hospital level at confirmed hypertension diagnosis, and comorbidities such as diabetes mellitus, hyperlipidemia, atrial fibrillation, hospitalization for heart failure, history of hospitalization for acute myocardial infarction, coronary artery disease, peripheral vascular disease, and chronic obstructive pulmonary disease. The detailed ICD codes are listed in **Supplementary Table 1**.

### Statistical Analysis

Baseline demographic and clinical characteristics of individuals with and without PA screening after a diagnosis of hypertension were compared using standardized differences (STD), with larger absolute STD values indicating substantial imbalance between groups. Inferential statistics were not applied due to the very large sample size (>7 million). We first calculated the overall screening rate as the proportion of patients with hypertension who underwent PA screening after their diagnosis of hypertension through the end of the database (December 31, 2023). Annual screening rates and diagnostic yields were then calculated as described in the study overview. Temporal trends in screening and diagnosis rates were assessed using the Cochran–Armitage trend test. To identify determinants of PA screening, we performed logistic regression including both patient-level covariates (age, sex, income, comorbidities, antihypertensive medication use) and system-level covariates (urbanization level of residence and hospital level). Because the same individual could appear in multiple years during the study period (2001–2022), the cluster effect within the same patient was accounted for using robust standard errors. All analyses were conducted using SAS version 9.4 (SAS Institute Inc., Cary, NC, USA). A two-sided *P* value <0.05 was considered statistically significant.

## Results

### Patient characteristics

During the study period from January 1, 2001, to December 31, 2022, a total of 8,039,134 patients with hypertension were identified from the NHIRD. After excluding 222,207 patients due to missing demographics, a final cohort of 7,816,927 patients was included for analysis (**Figure 1**). The baseline demographic and clinical characteristics of this nationwide cohort are presented in **Table 1**. The mean age at hypertension diagnosis was 56.8 ± 14.0 years, and 53.6% of the cohort were male. At baseline, the mean number of antihypertensive medication classes was 1.6 ± 0.9. Key high-risk phenotypes for PA were present, including hypokalemia in 1.6% of patients, early-onset hypertension in 11.3%, and resistant hypertension in 1.8%. The most common comorbidities at diagnosis of hypertension were diabetes mellitus (14.5%), dyslipidemia (13.3%), coronary artery disease (8.6%), and chronic kidney disease (5.0%).

**Figure 1.**
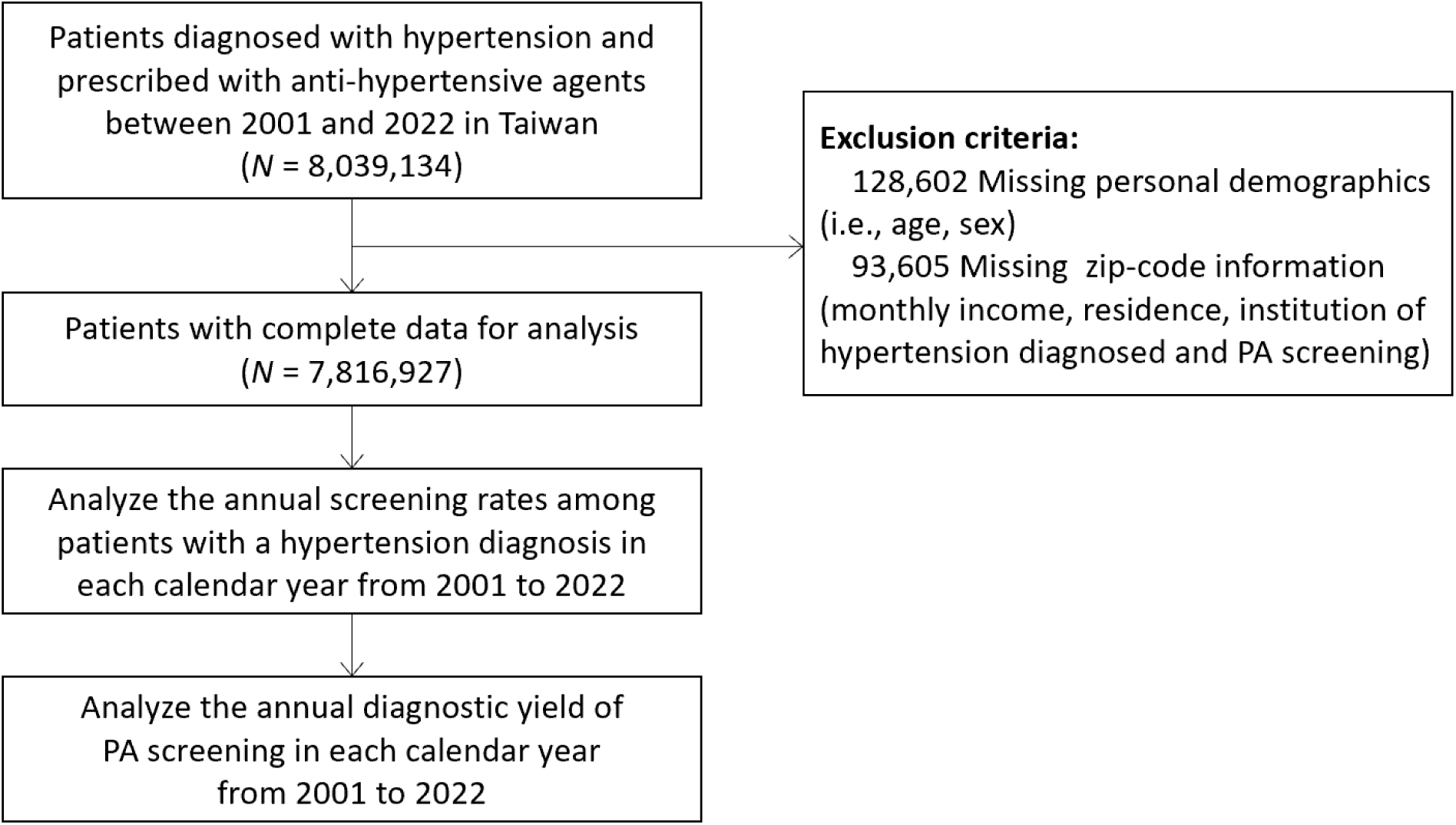
Study flowchart.

**Table 1.**
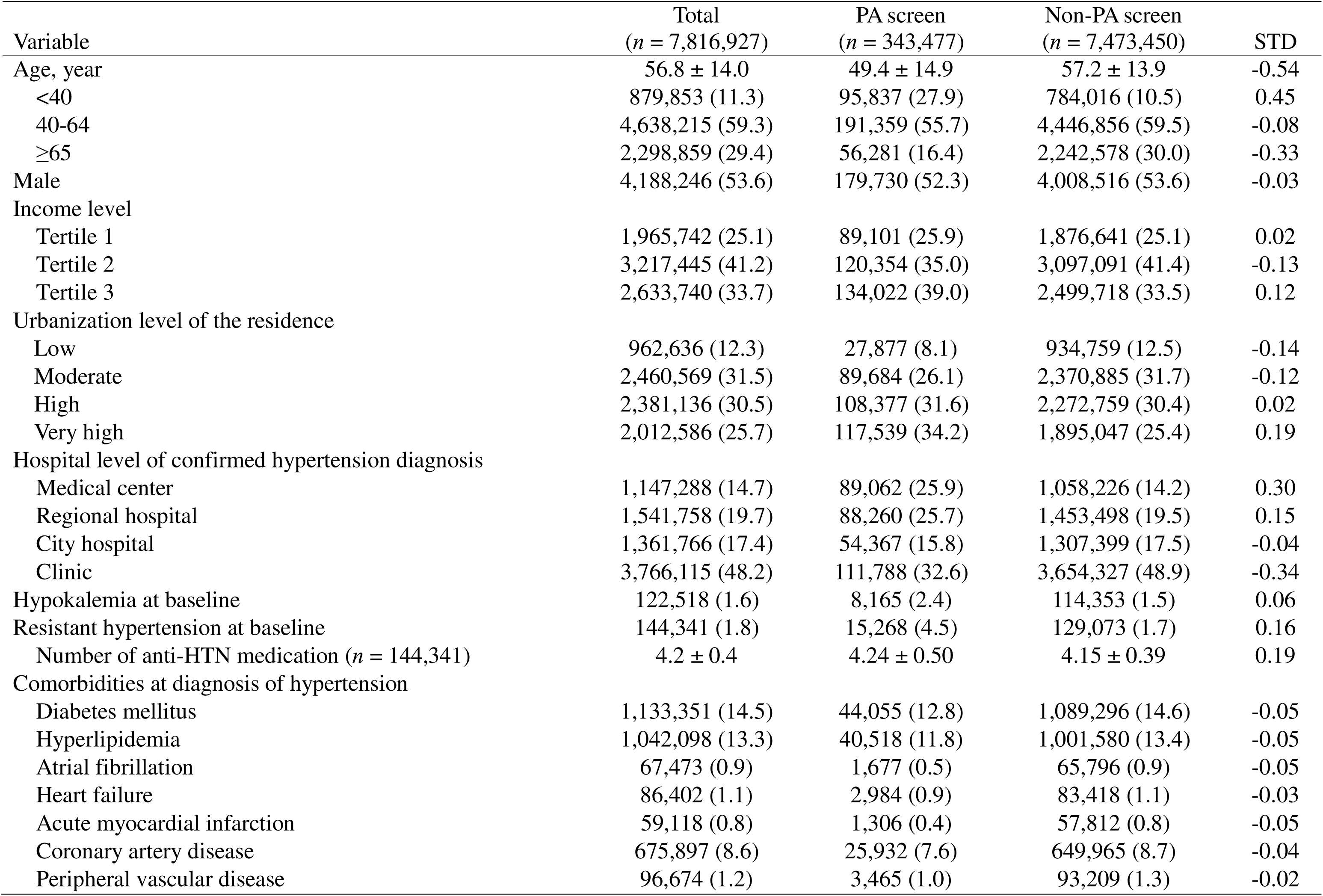
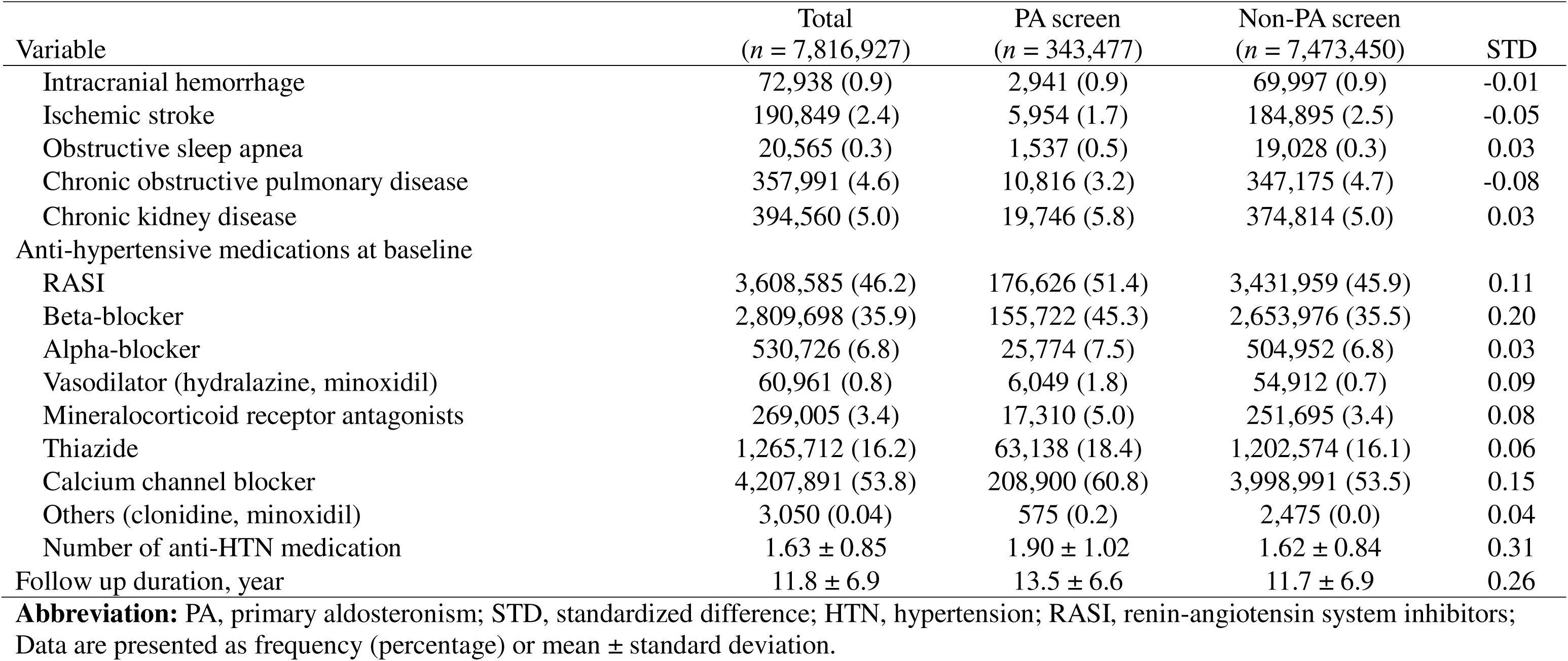
Baseline Demographic and Clinical Characteristics of Individuals with and without PA Screening after a Diagnosis of Hypertension.

| Variable | Total<br>(*n* = 7,816,927) | PA screen<br>(*n* = 343,477) | Non-PA screen<br>(*n* = 7,473,450) | STD |
| --- | --- | --- | --- | --- |
| Age, year | 56.8 ± 14.0 | 49.4 ± 14.9 | 57.2 ± 13.9 | -0.54 |
| <40 | 879,853 (11.3) | 95,837 (27.9) | 784,016 (10.5) | 0.45 |
| 40-64 | 4,638,215 (59.3) | 191,359 (55.7) | 4,446,856 (59.5) | -0.08 |
| ≥65 | 2,298,859 (29.4) | 56,281 (16.4) | 2,242,578 (30.0) | -0.33 |
| Male | 4,188,246 (53.6) | 179,730 (52.3) | 4,008,516 (53.6) | -0.03 |
| Income level |  |  |  |  |
| Tertile 1 | 1,965,742 (25.1) | 89,101 (25.9) | 1,876,641 (25.1) | 0.02 |
| Tertile 2 | 3,217,445 (41.2) | 120,354 (35.0) | 3,097,091 (41.4) | -0.13 |
| Tertile 3 | 2,633,740 (33.7) | 134,022 (39.0) | 2,499,718 (33.5) | 0.12 |
| Urbanization level of the residence |  |  |  |  |
| Low | 962,636 (12.3) | 27,877 (8.1) | 934,759 (12.5) | -0.14 |
| Moderate | 2,460,569 (31.5) | 89,684 (26.1) | 2,370,885 (31.7) | -0.12 |
| High | 2,381,136 (30.5) | 108,377 (31.6) | 2,272,759 (30.4) | 0.02 |
| Very high | 2,012,586 (25.7) | 117,539 (34.2) | 1,895,047 (25.4) | 0.19 |
| Hospital level of confirmed hypertension diagnosis |  |  |  |  |
| Medical center | 1,147,288 (14.7) | 89,062 (25.9) | 1,058,226 (14.2) | 0.30 |
| Regional hospital | 1,541,758 (19.7) | 88,260 (25.7) | 1,453,498 (19.5) | 0.15 |
| City hospital | 1,361,766 (17.4) | 54,367 (15.8) | 1,307,399 (17.5) | -0.04 |
| Clinic | 3,766,115 (48.2) | 111,788 (32.6) | 3,654,327 (48.9) | -0.34 |
| Hypokalemia at baseline | 122,518 (1.6) | 8,165 (2.4) | 114,353 (1.5) | 0.06 |
| Resistant hypertension at baseline | 144,341 (1.8) | 15,268 (4.5) | 129,073 (1.7) | 0.16 |
| Number of anti-HTN medication (*n* = 144,341) | 4.2 ± 0.4 | 4.24 ± 0.50 | 4.15 ± 0.39 | 0.19 |
| Comorbidities at diagnosis of hypertension |  |  |  |  |
| Diabetes mellitus | 1,133,351 (14.5) | 44,055 (12.8) | 1,089,296 (14.6) | -0.05 |
| Hyperlipidemia | 1,042,098 (13.3) | 40,518 (11.8) | 1,001,580 (13.4) | -0.05 |
| Atrial fibrillation | 67,473 (0.9) | 1,677 (0.5) | 65,796 (0.9) | -0.05 |
| Heart failure | 86,402 (1.1) | 2,984 (0.9) | 83,418 (1.1) | -0.03 |
| Acute myocardial infarction | 59,118 (0.8) | 1,306 (0.4) | 57,812 (0.8) | -0.05 |
| Coronary artery disease | 675,897 (8.6) | 25,932 (7.6) | 649,965 (8.7) | -0.04 |
| Peripheral vascular disease | 96,674 (1.2) | 3,465 (1.0) | 93,209 (1.3) | -0.02 |
| Intracranial hemorrhage | 72,938 (0.9) | 2,941 (0.9) | 69,997 (0.9) | -0.01 |
| Ischemic stroke | 190,849 (2.4) | 5,954 (1.7) | 184,895 (2.5) | -0.05 |
| Obstructive sleep apnea | 20,565 (0.3) | 1,537 (0.5) | 19,028 (0.3) | 0.03 |
| Chronic obstructive pulmonary disease | 357,991 (4.6) | 10,816 (3.2) | 347,175 (4.7) | -0.08 |
| Chronic kidney disease | 394,560 (5.0) | 19,746 (5.8) | 374,814 (5.0) | 0.03 |
| Anti-hypertensive medications at baseline |  |  |  |  |
| RASI | 3,608,585 (46.2) | 176,626 (51.4) | 3,431,959 (45.9) | 0.11 |
| Beta-blocker | 2,809,698 (35.9) | 155,722 (45.3) | 2,653,976 (35.5) | 0.20 |
| Alpha-blocker | 530,726 (6.8) | 25,774 (7.5) | 504,952 (6.8) | 0.03 |
| Vasodilator (hydralazine, minoxidil) | 60,961 (0.8) | 6,049 (1.8) | 54,912 (0.7) | 0.09 |
| Mineralocorticoid receptor antagonists | 269,005 (3.4) | 17,310 (5.0) | 251,695 (3.4) | 0.08 |
| Thiazide | 1,265,712 (16.2) | 63,138 (18.4) | 1,202,574 (16.1) | 0.06 |
| Calcium channel blocker | 4,207,891 (53.8) | 208,900 (60.8) | 3,998,991 (53.5) | 0.15 |
| Others (clonidine, minoxidil) | 3,050 (0.04) | 575 (0.2) | 2,475 (0.0) | 0.04 |
| Number of anti-HTN medication | 1.63 ± 0.85 | 1.90 ± 1.02 | 1.62 ± 0.84 | 0.31 |
| Follow up duration, year | 11.8 ± 6.9 | 13.5 ± 6.6 | 11.7 ± 6.9 | 0.26 |

**Abbreviation:** PA, primary aldosteronism; STD, standardized difference; HTN, hypertension; RASI, renin-angiotensin system inhibitors; Data are presented as frequency (percentage) or mean ± standard deviation.

### Temporal trends in overall PA screening

During 2001–2022, a total of 64 million records were extracted, representing the annual number of individuals diagnosed with hypertension, including both prevalent and incident cases (**Figure 1**). As illustrated in **Figure 2A** and **Supplementary Table 2**, the annual screening rate for PA among all patients with hypertension demonstrated a significant, nearly threefold increase over the 22-year study period. The rate rose from 0.26% in 2001 to a peak of 0.75% in 2022 (P for trend < 0.001). Despite this upward trend, the absolute annual screening rate remained markedly low throughout the two decades (0.55%; 353,971 out of 64,432,085).

**Figure 2.**
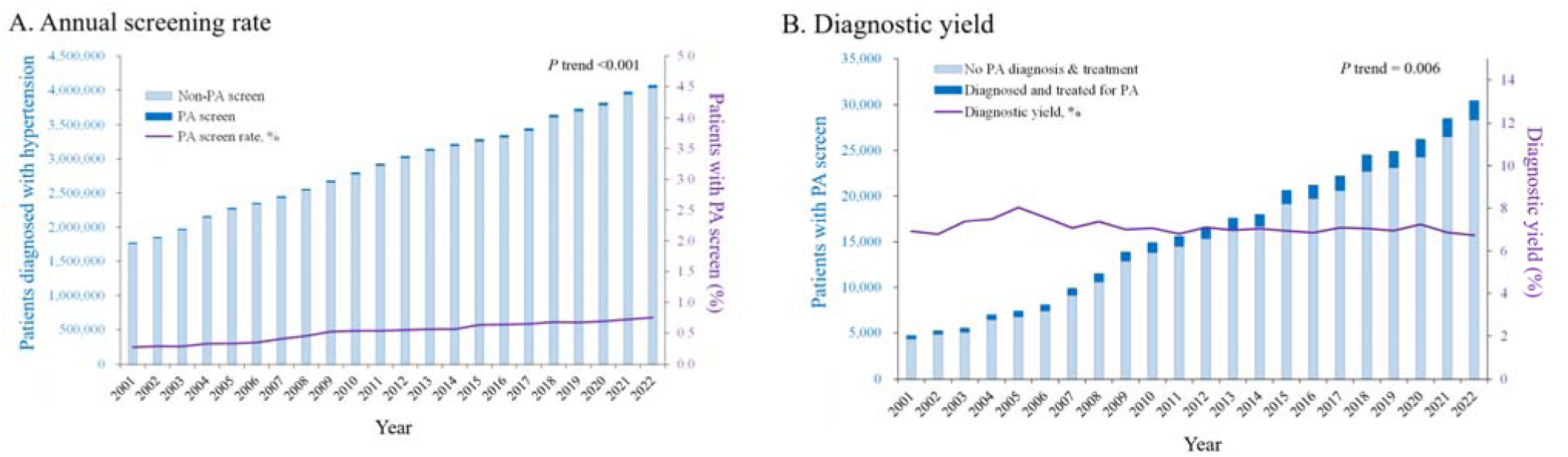
Annual screening rate and new diagnoses cases for primary aldosteronism. (A) The figure illustrates the temporal trend in the annual screening rate for primary aldosteronism. The screening rate is expressed as a percentage of the total living hypertensive population each year. (B) This figure shows the annual number of newly confirmed diagnoses of primary aldosteronism resulting in treatment and the diagnostic yield.

### Screening rates within high-risk subgroups

Analysis of temporal trends among high-risk subgroups demonstrated that while screening rates increased across all evaluated groups, both the magnitude of the increase and the final rates in 2022 varied substantially (**Figure 3, Supplementary Table 3-5**). The most pronounced increases were observed among patients with hypokalemia (from 2.3% in 2001 to 3.6% in 2022), early-onset hypertension (from 1.9% to 3.2%), obstructive sleep apnea (from 0.3% to 1.9%) and resistant hypertension (from 0.6% to 1.0%). Despite these improvements, screening rates remained critically low in precisely those subgroups that guidelines prioritize for testing(31). Furthermore, annual screening rates for PA among patients with other comorbidities, including chronic kidney disease, ischemic stroke, and hemorrhagic stroke, were all less than 1% in 2022. All of these upward trends were statistically significant (*P* for trend < 0.001 for all subgroups).

**Figure 3.**
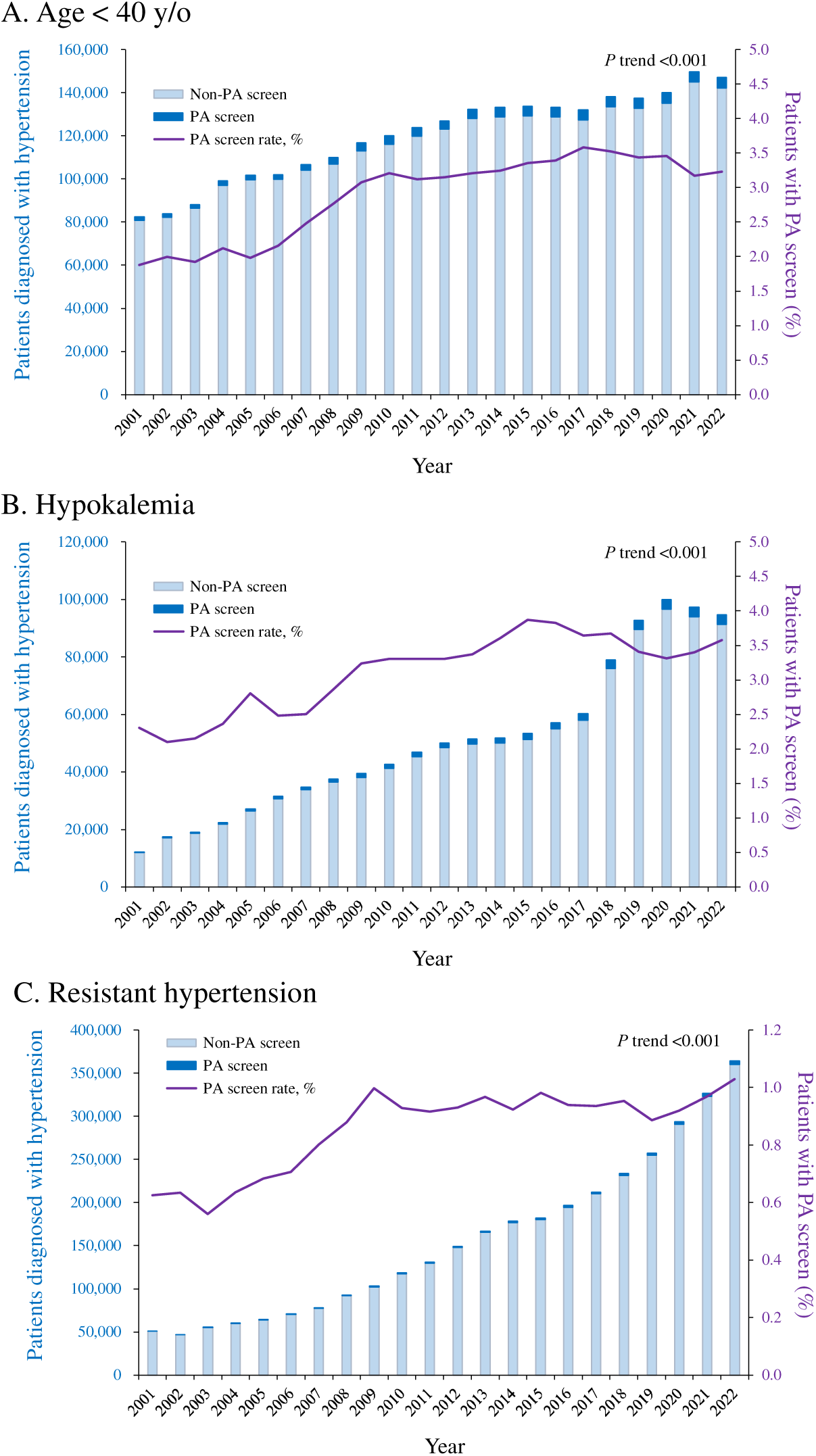
Annual screening rate for primary aldosteronism in high-risk population. The figure illustrates the temporal trend in the annual screening rate for primary aldosteronism among different high-risk population including (A) early onset hypertension, (B) hypertension with hypokalemia, (C) resistant hypertension. The screening rate is expressed as a percentage of the total living hypertensive population each year.

### Factors Associated with Screening for PA

The results of factors associated with receiving PA screening are presented in **Figure 4**. Both patient- and system-level factors were significant factors. At the patient level, younger age (age <40 years vs. ≥65 years: odds ratio [OR] 8.95, 95% confidence interval [CI] 8.86–9.03) and the presence of hypokalemia (OR 6.82, 95% CI 6.74–6.89) were the strongest determinants of screening. Other clinical conditions including resistant hypertension (OR 1.74, 95% CI 1.72–1.76), chronic kidney disease, obstructive sleep apnea, and a history of intracranial hemorrhage were also significantly associated with an increased likelihood of screening, whereas a history of ischemic stroke was not. Patients diagnosed in more recent years were more likely to be screened. At the system level, residing in highly urbanized areas and receiving initial care at a medical center were associated with greater screening likelihood. In contrast, patient income level was not significantly related to PA screening.

**Figure 4.**
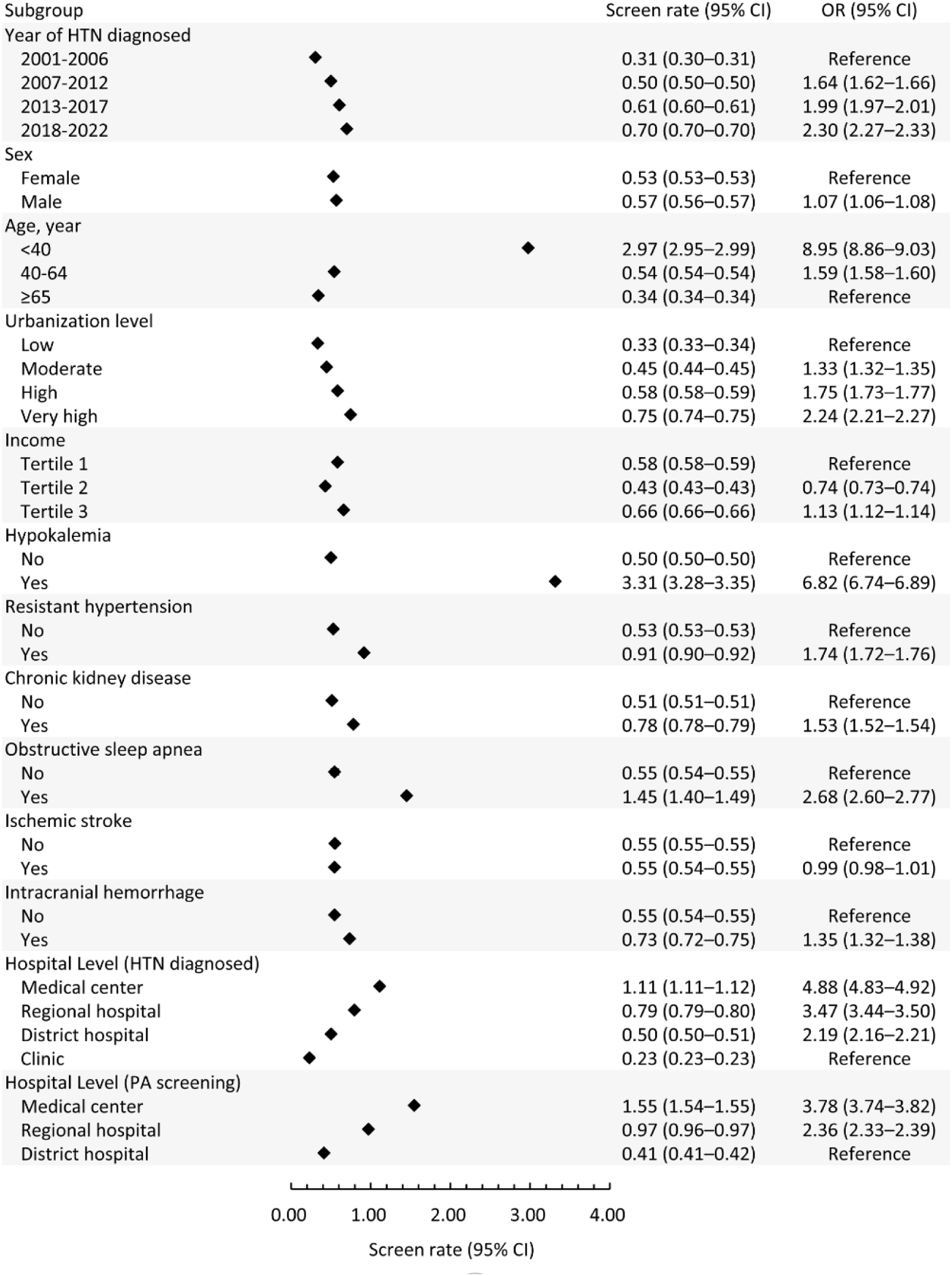
Factors Associated with Screening for PA. This forest plot displays the odds ratio and 95% confidence intervals for factors associated with the likelihood of receiving PA screening.

### Trends in PA diagnosis and diagnostic yield

Over the 22-year study period, the absolute number of new PA diagnoses per year increased steadily, rising from 324 cases (6.9%) in 2001 to 2,051 cases (6.7%) in 2022 (**Figure 2B** and **Supplementary Table 2**). In contrast to the rising number of screenings, the diagnostic yield, defined as the proportion of screened patients who received a confirmed PA diagnosis and PA treatment, remained relatively stable. The diagnostic yield fluctuated between 8.1% and 6.8% throughout the study period, with a modest but significant temporal trend of decline observed (*P* for trend = 0.006)

### PA diagnosis and diagnostic yield within high-risk subgroups

The diagnostic yield of PA varied considerably across different high-risk subgroups (**Supplementary Table 3-5**). The highest diagnostic yield was observed in patients with hypokalemia (19.2%), followed by those with resistant hypertension (11.7%), obstructive sleep apnea (9.8%), a history of intracranial hemorrhage (8.0%), chronic kidney disease (6.3%), and a history of ischemic stroke (5.8%). When stratified by age, the diagnostic yield was highest in the 40-65 age group (9.1%), compared to both the < 40 and ≥ 65 age groups (5.0% for each). A modest but statistically significant temporal decline in diagnostic yield was observed specifically within all age subgroups, as well as in patients with hypokalemia, resistant hypertension, and chronic kidney disease (P for trend < 0.001).

## Discussion

In this nationwide, population-based study spanning two decades, we demonstrated the evolving landscape of PA screening and diagnosis in Taiwan. While the annual screening rate for PA tripled during this period, it remained critically low at less than 1% among patients with hypertension. This under-screening was particularly pronounced in guideline-recommended high-risk subgroups, revealing a significant care gap. Notably, however, despite a steady rise in screening, the diagnostic yield remained remarkably stable, suggesting that increased test utilization did not translate into improved case detection within the current clinical pathway. To our knowledge, this is the first study to provide long-term, nationwide data on these trends in Taiwan.

Our primary finding of a threefold increase in annual screening rates, from 0.26% to 0.75%, suggests a growing, albeit slow, awareness of PA among clinicians. This trend likely reflects the cumulative effect of guideline publications and educational efforts by professional societies over the past two decades. However, this modest progress is overshadowed by the persistently low absolute screening rate. Importantly, by evaluating the entire hypertensive population rather than only high-risk subgroups, our study establishes a unique nationwide benchmark: fewer than 1% of hypertensive patients are screened annually. This provides a unique nationwide benchmark that directly contrasts with the new guideline recommendation of universal testing for all hypertensive patients. When placed in an international context, our findings echo similarly low screening rates observed in high-risk Western cohort. For instance, a large study of U.S. veterans reported a similarly low screening rate of 1.6% in patients with resistant hypertension(24), while a nationwide study in Sweden also concluded that PA remains profoundly underdiagnosed(32). Turcu et al. reported that in a tertiary referral academic center, the screening rate for PA was only 3.4%, even among patients who met the guideline-recommended criteria for screening(25). Similarly, a recent multicenter study by Kositanurit et al. also reported stagnant temporal trends in PA screening among high-risk patients(33). In addition, a prior population-based cohort study by Liu et al. in Alberta, Canada, reported similarly low overall PA screening rates (0.7%) across the hypertensive population; however, screening rates were higher (1.7%) in the geographic region served by a specialized endocrine hypertension program(34). Taken together, these results highlight a strikingly consistent implementation gap across diverse healthcare systems, suggesting that the root causes of under-screening such as provider education, diagnostic complexity, and inertia in clinical practice are universal rather than system-specific. Our data add to this literature by providing the first nationwide “all-comers” perspective, quantifying just how far current practice must advance to achieve the newly proposed standard of universal PA screening.

The underperformance of screening was also evident in the populations who stand to benefit the most. The fact that only 0.9% of patients with resistant hypertension, 3.0% of those with early-onset hypertension and 3.3% of those with hypertension and hypokalemia were screened in this study is particularly concerning, as these groups have the highest reported prevalence of PA. This persistent clinical inertia likely reflects several barriers identified in previous literature, including a lack of provider familiarity with guidelines in different subspecialties, pre-analytical sample preparation and laboratory infrastructure constraints, the perceived complexity of managing and withdrawing potentially interfering antihypertensive medications, knowledge gaps in interpreting screening results, the laborious nature of the multi-step diagnostic pathway for PA, and limited access to specialists for interpretation and management(35–41). This may also reflect a persistent, outdated view of PA as a rare disease defined only by classic phenotypes like resistant hypertension with hypokalemia, overlooking the broader spectrum of renin-independent aldosterone production now known to be common(42).

Another interesting finding of our study is that hypertensive patients who first present to academic medical centers are more likely to receive screening. This suggests that the workup for PA is still largely driven by specialists, likely hypertension specialists, endocrinologists, nephrologists and cardiologist in tertiary care settings. While specialist-driven care ensures expertise, it also implies that a vast majority of hypertensive patients managed in regional, district, or primary care settings may have limited access to screening, creating a potential issue of health inequity and missed opportunities for diagnosis which urgently need further intervention(36,37). To address this, Libianto et al. demonstrated that empowering and educating primary care physicians to screen for PA is a highly effective approach(43). More recently, Brown et al. proposed a “direct-to-patient” testing program that simplifies protocols and directly engages patients; such a model could help decentralize screening and improve detection rates outside of major academic hospitals(44). Importantly, the 2025 Endocrine society guideline also emphasizes simplified, pragmatic recommendations for screening and diagnosing PA(22).

Perhaps the most counterintuitive finding of our study is the stable, low diagnostic yield of approximately 7-8%, and even a modest but statistically significant decline over the two-decade period. This slight decline may paradoxically reflect a positive trend: as physician awareness and screening slowly expanded from only the most classic cases to a broader, less phenotypically severe high-risk population, the overall diagnostic yield became diluted. However, this dilution effect does not explain the larger issue of why the yield was so low to begin with. Given that screening was largely confined to high-risk subgroups where PA prevalence is estimated to be as high as 20%(42,45,46), a much higher yield would be expected. This discrepancy strongly suggests that the bottleneck is not only in selecting the right patients for screening, but also in the complex, multi-step diagnostic pathway that follows a positive initial screen. Factors such as the requirement for medication withdrawal, a significant barrier for clinicians, and the high false-negative rates of confirmatory tests themselves likely lead to a substantial number of true-positive cases being lost during the workup(39,47).

Several factors within the current diagnostic algorithm contribute to this phenomenon. First, the recommended screening cutoff for the aldosterone-to-renin ratio in this study region is relatively strict (ARR > 30 to 35 ng/dL per ng/mL per hour), which may cause false negatives at the initial stage(18,48). Second, the subsequent confirmatory process is complex and requires medication adjustments, which may deter non-specialist physicians from proceeding to a final diagnosis(21,48–50). Compounding this issue, recent evidence has raised concerns about the high false-negative rates of the confirmatory aldosterone suppression tests themselves(39,47,51,52). Importantly, these systemic barriers are directly addressed by the 2025 Endocrine Society guideline, which proposes removing the routine requirement for aldosterone suppression tests, applying more permissive diagnostic cutoffs, and allowing screening without discontinuing most antihypertensive medications(22). Collectively, these changes are expected to simplify the diagnostic process, empower primary care physicians to initiate screening, and ultimately increase the diagnostic yield, broadening detection beyond only those with the most extreme disease presentations.

This profound under-screening represents a significant missed opportunity, not only for improving clinical outcomes but also for healthcare efficiency. A cost-effectiveness analysis by Lubitz et al. demonstrated that screening for PA in patients with resistant hypertension, followed by appropriate surgical or medical treatment, is a cost-effective strategy(53). The upfront costs of screening are offset by long-term gains in quality-adjusted life years and reductions in cardiovascular events. Our findings, which quantify the scale of under-screening, therefore imply that the healthcare system is failing to capitalize on these potential long-term benefits.

This study has several limitations inherent to its retrospective design using an administrative claims database. First, diagnoses are based on ICD codes and may be subject to misclassification. Second, we lack granular clinical data, such as specific blood pressure values, serum potassium levels, and the actual results of renin and aldosterone tests, which prevents a more detailed analysis of the screened population’s characteristics. Third, our operational definition for a confirmed PA diagnosis, while robust for this study type, cannot replace a formal diagnosis based on guideline-recommended testing and results interpretation. Moreover, because our definition required both an ICD code and the initiation of aldosterone-targeted treatment, it may have underestimated the true number of confirmed cases. Finally, as a retrospective analysis of claims data, we could not ascertain the physician’s specific motivation for ordering tests or adherence to pre-test protocols or their interpretation of the results.

## Conclusion

In conclusion, this study reveals that despite progress over the last two decades, PA remains profoundly underdiagnosed on a nationwide scale, with a major implementation gap between guideline recommendations and clinical practice, in Taiwan as in other countries around the world and despite universal insurance coverage. Addressing this long-standing care gap may require a fundamental shift in the diagnostic and therapeutic paradigm for PA. This could involve adopting permissive screening criteria to increase sensitivity at the initial stage. Furthermore, simplifying the diagnostic pathway by waiving routine aldosterone suppression testing as current guideline suggested could empower more clinicians to make a diagnosis(22). Ultimately, a more liberal implementation of MRA therapy for patients with clear evidence of renin-independent aldosterone production may be warranted, even if they do not meet the full, strict criteria for a confirmed PA diagnosis(38,54). Our findings also highlight the urgent need for system-level interventions to decentralize screening and empower primary care, which may ultimately reduce the cardiovascular burden of PA. The field of testing, diagnosis and management of PA is currently undergoing a significant paradigm shift. Therefore, future prospective studies will be crucial to evaluate the impact of these changes on clinical practice and patient outcomes.

## Data Availability

All data produced in the present study are available upon reasonable request to the corresponding author.

## Acknowledgements

The authors thank the staff of the Eighth and Second Core Lab in the Department of Medical Research at National Taiwan University Hospital for technical support. We also thank all members of the TAIPAI Study Group (https://doi.org/10.6084/m9.figshare.21669929.v7) for help during the study. The authors also thank Alfred Hsing-Fen Lin, Zoe Ya-Zhu Syu, and Winnie Su-Hui Wu, who served in Raising Statistics Consultant Inc., for their assistance with statistical analysis.

---

**Supplementary Table 1.** Diagnostic Codes for Diseases Used in This Study.

| Disease | ICD-9-CM diagnostic code | ICD-10-CM diagnostic code |
| --- | --- | --- |
| Hypertension | 401.xx-405.xx | I10-I15, N262 |
| Primary aldosteronism | 255.1 | E26.01, E26.02, E26.09, E26.89, E26.9 |
| Hypokalemia | 276.8 | E87.6 |
| Diabetes mellitus | 250.x | E08-E13 |
| Hyperlipidemia | 272.0-272.4 | E77, E780, E781, E782, E783, E784, E785, E786, E881, E753, E755, E882, E756, E789, E7521, E7522, E7524, E7130, E7879, E7881, E7889, E8889, E7870 |
| Atrial fibrillation | 427.31 | I480-I482, I4891 |
| Heart failure | 428.x | I50 |
| Acute myocardial infarction | 410.x | I21-I22 |
| Myocardial infarction | 410.x, 412.x | I21-I22, I25.2 |
| Coronary artery disease | 410.x-414.x | I20-I22, I24, I25 |
| Peripheral vascular disease | 440.0x, 440.2x, 440.3x, 440.8x, 440.9x, 443.x, 444.0x, 444.22, 444.8x, 447.8x, 447.9x | I70, I71, I73, I75, I771, I790, I791, I792, I773, I779, I798, K551, K558, K559, Z958, Z959, I743, I744, I745, I748, I740, I7789 |
| Intracranial hemorrhage | 430.x-432.x | I60-I62 |
| Ischemic stroke | 433.x-437.x | G450, G451, G452, G454, G458, G459, G460, G461, G462, G463, G464, G465, G466, G467, G468, I6300, I63011, I63012, I63019, I6302, I6303, I6309, I6310, I63111, I63112, I63119, I6312, I6313, I6319, I6320, I63211, I63212, I63219, I6322, I6323, I6329, I6330, I6331, I63311, I63312, I63319, I63321, I63322, I63329, I6333, I63331, I63332, I63339, I6334, I63341, I63342, I63349, I6339, I6340, I6341, I63411, I63412, I63419, I6342, I63421, I63422, I63429, I63431, I63432, I63439, I63441, I63442, I63449, I6349, I6350, I63511, I63512, I63519, I63521, I63522, I63529, I63531, I63532, I63539, I63541, I63542, I63549, I6359, I6359, I636, I638, I639, I650, I651, I6521, I6522, I6523, I6529, I658, I659, I66, I670, I671, I672, I674, I675, I676, I677, I6781, I6782, |
|  |  | I6789, I679, I680, I682, I688 |
| Obstructive sleep apnea | 327.23, 780.51, 780.53, 780.57 | G47.30, G47.31, G47.33, G47.37, G47.39 |
| Chronic obstructive pulmonary disease | 491.x, 492.x, 496.x | J41-J44 |
| Chronic kidney disease | 580.x–589.x, 403.x–404.x, 016.0x,<br>095.4x, 236.9x, 250.4x, 274.1x,<br>442.1x, 447.3x, 440.1x, 572.4x,<br>642.1x, 646.2x, 753.1x, 283.11,<br>403.01, 404.02, 446.21 | A1811, D593, E102, E112, E132, I12, I13, K767,<br>M103, M310, N00, N01, N02, N03, N04, N05,<br>N06, N07, N08, N14, N150, N158, N159, N16,<br>N171, N172, N18, N19, N200, N25, N261, N269,<br>N27, Q61 |

**Abbreviation:** ICD-9-CM, international classification of diseases, ninth revision, clinical modification; ICD-10-CM, international classification of diseases, tenth revision, clinical modification.

**Supplementary Table 2.** Annual Counts and Proportions of PA Screening and PA Diagnosis among Hypertensive Individuals.

| Year | No. of individuals diagnosed with hypertension | No. of individuals with hypertension receiving PA screening | No. of individuals confirmed PA after PA screening* |
| --- | --- | --- | --- |
| 2001 | 1,768,758 | 4,680 (0.26) | 324 (6.9) |
| 2002 | 1,850,021 | 5,203 (0.28) | 353 (6.8) |
| 2003 | 1,971,440 | 5,501 (0.28) | 406 (7.4) |
| 2004 | 2,154,944 | 6,979 (0.32) | 522 (7.5) |
| 2005 | 2,275,101 | 7,386 (0.32) | 593 (8.0) |
| 2006 | 2,349,912 | 8,049 (0.34) | 609 (7.6) |
| 2007 | 2,444,943 | 9,849 (0.40) | 697 (7.1) |
| 2008 | 2,554,194 | 11,473 (0.45) | 846 (7.4) |
| 2009 | 2,676,987 | 13,869 (0.52) | 971 (7.0) |
| 2010 | 2,796,042 | 14,887 (0.53) | 1,052 (7.1) |
| 2011 | 2,923,219 | 15,579 (0.53) | 1,059 (6.8) |
| 2012 | 3,034,036 | 16,543 (0.55) | 1,176 (7.1) |
| 2013 | 3,140,809 | 17,553 (0.56) | 1,224 (7.0) |
| 2014 | 3,211,652 | 17,967 (0.56) | 1,266 (7.0) |
| 2015 | 3,280,623 | 20,600 (0.63) | 1,429 (6.9) |
| 2016 | 3,340,843 | 21,186 (0.63) | 1,452 (6.9) |
| 2017 | 3,439,194 | 22,185 (0.65) | 1,573 (7.1) |
| 2018 | 3,639,679 | 24,471 (0.67) | 1,725 (7.0) |
| 2019 | 3,727,772 | 24,878 (0.67) | 1,728 (6.9) |
| 2020 | 3,817,387 | 26,203 (0.69) | 1,899 (7.2) |
| 2021 | 3,969,164 | 28,491 (0.72) | 1,955 (6.9) |
| 2022 | 4,065,365 | 30,439 (0.75) | 2,051 (6.7) |
| **Total** | **64,432,085** | **353,971 (0.55)** | **24,910 (7.0)** |
| *P* for trend | - | <0.001 | 0.006 |

**Abbreviation:** PA, primary aldosteronism;
\* The denominator was the number of patients with hypertension who underwent PA screening in that calendar year.

**Supplementary Table 3.** Annual Counts and Proportions of PA Screening and PA Diagnosis among Hypertensive Individuals among Age Groups.

| Year | Age <40 years |  |  | Age 40-64 years |  |  | Age ≥65 years |  |  |
| --- | --- | --- | --- | --- | --- | --- | --- | --- | --- |
|  | Hypertension | PA screening | Confirmed PA* | Hypertension | PA screening | Confirmed PA* | Hypertension | PA screening | Confirmed PA* |
| 2001 | 82,369 | 1,549 (1.9) | 115 (7.4) | 875,899 | 2,155 (0.25) | 179 (8.3) | 810,490 | 976 (0.12) | 30 (3.1) |
| 2002 | 83,775 | 1,674 (2.0) | 109 (6.5) | 916,910 | 2,446 (0.27) | 208 (8.5) | 849,336 | 1,083 (0.13) | 36 (3.3) |
| 2003 | 88,106 | 1,693 (1.9) | 112 (6.6) | 982,430 | 2,627 (0.27) | 247 (9.4) | 900,904 | 1,181 (0.13) | 47 (4.0) |
| 2004 | 99,157 | 2,101 (2.1) | 133 (6.3) | 1,086,309 | 3,276 (0.30) | 320 (9.8) | 969,478 | 1,602 (0.17) | 69 (4.3) |
| 2005 | 101,689 | 2,014 (2.0) | 129 (6.4) | 1,150,662 | 3,419 (0.30) | 387 (11.3) | 1,022,750 | 1,953 (0.19) | 77 (3.9) |
| 2006 | 102,045 | 2,203 (2.2) | 139 (6.3) | 1,185,409 | 3,664 (0.31) | 369 (10.1) | 1,062,458 | 2,182 (0.21) | 101 (4.6) |
| 2007 | 106,735 | 2,651 (2.5) | 143 (5.4) | 1,234,459 | 4,662 (0.38) | 455 (9.8) | 1,103,749 | 2,536 (0.23) | 99 (3.9) |
| 2008 | 109,909 | 3,041 (2.8) | 175 (5.8) | 1,294,901 | 5,479 (0.42) | 565 (10.3) | 1,149,384 | 2,953 (0.26) | 106 (3.6) |
| 2009 | 116,640 | 3,585 (3.1) | 183 (5.1) | 1,369,329 | 6,610 (0.48) | 654 (9.9) | 1,191,018 | 3,674 (0.31) | 134 (3.6) |
| 2010 | 119,970 | 3,852 (3.2) | 214 (5.6) | 1,438,615 | 7,164 (0.50) | 671 (9.4) | 1,237,457 | 3,871 (0.31) | 167 (4.3) |
| 2011 | 123,670 | 3,858 (3.1) | 199 (5.2) | 1,522,875 | 7,679 (0.50) | 692 (9.0) | 1,276,674 | 4,042 (0.32) | 168 (4.2) |
| 2012 | 126,996 | 4,002 (3.2) | 190 (4.7) | 1,588,375 | 8,131 (0.51) | 794 (9.8) | 1,318,665 | 4,410 (0.33) | 192 (4.4) |
| 2013 | 132,330 | 4,250 (3.2) | 220 (5.2) | 1,643,212 | 8,700 (0.53) | 776 (8.9) | 1,365,267 | 4,603 (0.34) | 228 (5.0) |
| 2014 | 133,139 | 4,324 (3.2) | 218 (5.0) | 1,666,957 | 8,797 (0.53) | 833 (9.5) | 1,411,556 | 4,846 (0.34) | 215 (4.4) |
| 2015 | 133,773 | 4,483 (3.4) | 199 (4.4) | 1,680,962 | 10,179 (0.61) | 936 (9.2) | 1,465,888 | 5,938 (0.41) | 294 (5.0) |
| 2016 | 133,167 | 4,516 (3.4) | 208 (4.6) | 1,692,046 | 10,802 (0.64) | 972 (9.0) | 1,515,630 | 5,868 (0.39) | 272 (4.6) |
| 2017 | 132,050 | 4,727 (3.6) | 204 (4.3) | 1,704,518 | 11,167 (0.66) | 1,041 (9.3) | 1,602,626 | 6,291 (0.39) | 328 (5.2) |
| 2018 | 138,173 | 4,868 (3.5) | 224 (4.6) | 1,769,040 | 12,428 (0.70) | 1,080 (8.7) | 1,732,466 | 7,175 (0.41) | 421 (5.9) |
| 2019 | 137,506 | 4,726 (3.4) | 193 (4.1) | 1,788,067 | 12,613 (0.71) | 1,136 (9.0) | 1,802,199 | 7,539 (0.42) | 399 (5.3) |
| 2020 | 139,957 | 4,839 (3.5) | 229 (4.7) | 1,807,167 | 13,051 (0.72) | 1,163 (8.9) | 1,870,263 | 8,313 (0.44) | 507 (6.1) |
| 2021 | 149,586 | 4,749 (3.2) | 213 (4.5) | 1,862,264 | 14,078 (0.76) | 1,184 (8.4) | 1,957,314 | 9,664 (0.49) | 558 (5.8) |
| 2022 | 146,956 | 4,743 (3.2) | 202 (4.3) | 1,883,119 | 15,148 (0.80) | 1,246 (8.2) | 2,035,290 | 10,548 (0.52) | 603 (5.7) |
| **Total** | **2,637,698** | **78,448 (3.0)** | **3,951 (5.0)** | **32,143,525** | **174,275 (0.54)** | **15,908 (9.1)** | **29,650,862** | **101,248 (0.34)** | **5,051 (5.0)** |
| *P* for trend | - | <0.001 | <0.001 | - | <0.001 | <0.001 | - | <0.001 | <0.001 |

**Abbreviation:** PA, primary aldosteronism;
\* The denominator was the number of patients with hypertension who underwent PA screening in that calendar year.

**Supplementary Table 4.** Annual Counts and Proportions of PA Screening and PA Diagnosis among Hypertensive Individuals among Potentially High-Risk Subgroups: Hypokalemia, Resistant Hypertension and Chronic Kidney Disease.

| Year | Hypokalemia |  |  | Resistant hypertension |  |  | Chronic kidney disease |  |  |
| --- | --- | --- | --- | --- | --- | --- | --- | --- | --- |
|  | Hypertension | PA screening | Confirmed PA* | Hypertension | PA screening | Confirmed PA* | Hypertension | PA screening | Confirmed PA* |
| 2001 | 12,274 | 283 (2.3) | 66 (23.3) | 51,073 | 319 (0.62) | 44 (13.8) | 199,143 | 1,022 (0.51) | 41 (4.0) |
| 2002 | 17,505 | 368 (2.1) | 109 (29.6) | 47,089 | 299 (0.63) | 31 (10.4) | 206,551 | 1,148 (0.56) | 57 (5.0) |
| 2003 | 19,166 | 413 (2.2) | 94 (22.8) | 55,475 | 311 (0.56) | 49 (15.8) | 212,963 | 1,032 (0.48) | 80 (7.8) |
| 2004 | 22,414 | 530 (2.4) | 134 (25.3) | 60,391 | 384 (0.64) | 52 (13.5) | 221,468 | 1,330 (0.60) | 93 (7.0) |
| 2005 | 27,264 | 764 (2.8) | 167 (21.9) | 64,621 | 442 (0.68) | 66 (14.9) | 229,664 | 1,441 (0.63) | 92 (6.4) |
| 2006 | 31,535 | 784 (2.5) | 162 (20.7) | 70,848 | 501 (0.71) | 70 (14.0) | 228,677 | 1,592 (0.70) | 89 (5.6) |
| 2007 | 34,749 | 870 (2.5) | 187 (21.5) | 77,967 | 625 (0.80) | 63 (10.1) | 232,055 | 1,838 (0.79) | 79 (4.3) |
| 2008 | 37,531 | 1,074 (2.9) | 203 (18.9) | 92,667 | 814 (0.88) | 103 (12.7) | 240,904 | 2,137 (0.89) | 108 (5.1) |
| 2009 | 39,459 | 1,277 (3.2) | 262 (20.5) | 103,427 | 1,031 (1.00) | 112 (10.9) | 246,761 | 2,465 (1.00) | 129 (5.2) |
| 2010 | 42,633 | 1,409 (3.3) | 273 (19.4) | 118,432 | 1,099 (0.93) | 133 (12.1) | 254,845 | 2,496 (0.98) | 150 (6.0) |
| 2011 | 46,882 | 1,550 (3.3) | 277 (17.9) | 130,725 | 1,197 (0.92) | 117 (9.8) | 267,280 | 2,556 (0.96) | 128 (5.0) |
| 2012 | 50,186 | 1,657 (3.3) | 333 (20.1) | 149,293 | 1,388 (0.93) | 130 (9.4) | 318,871 | 2,794 (0.88) | 163 (5.8) |
| 2013 | 51,471 | 1,735 (3.4) | 332 (19.1) | 166,884 | 1,614 (0.97) | 157 (9.7) | 365,321 | 3,002 (0.82) | 186 (6.2) |
| 2014 | 51,878 | 1,869 (3.6) | 341 (18.2) | 178,768 | 1,649 (0.92) | 188 (11.4) | 403,414 | 3,193 (0.79) | 200 (6.3) |
| 2015 | 53,435 | 2,068 (3.9) | 399 (19.3) | 181,904 | 1,785 (0.98) | 199 (11.1) | 440,069 | 3,838 (0.87) | 227 (5.9) |
| 2016 | 57,093 | 2,184 (3.8) | 397 (18.2) | 196,471 | 1,846 (0.94) | 200 (10.8) | 475,602 | 3,730 (0.78) | 233 (6.2) |
| 2017 | 60,278 | 2,196 (3.6) | 437 (19.9) | 212,119 | 1,983 (0.93) | 237 (12.0) | 536,668 | 4,120 (0.77) | 285 (6.9) |
| 2018 | 78,874 | 2,896 (3.7) | 553 (19.1) | 233,875 | 2,230 (0.95) | 260 (11.7) | 640,842 | 5,021 (0.78) | 335 (6.7) |
| 2019 | 92,727 | 3,159 (3.4) | 586 (18.6) | 257,289 | 2,282 (0.89) | 252 (11.0) | 700,443 | 5,167 (0.74) | 348 (6.7) |
| 2020 | 99,855 | 3,311 (3.3) | 652 (19.7) | 293,544 | 2,700 (0.92) | 351 (13.0) | 735,359 | 5,523 (0.75) | 395 (7.2) |
| 2021 | 97,333 | 3,312 (3.4) | 586 (17.7) | 326,316 | 3,162 (0.97) | 396 (12.5) | 767,011 | 5,964 (0.78) | 430 (7.2) |
| 2022 | 94,685 | 3,385 (3.6) | 573 (16.9) | 363,869 | 3,745 (1.03) | 472 (12.6) | 794,627 | 6,740 (0.85) | 463 (6.9) |
| **Total** | **1,119,227** | **37,094 (3.3)** | **7,123 (19.2)** | **3,433,047** | **31,406 (0.91)** | **3,682 (11.7)** | **8,718,538** | **68,149 (0.78)** | **4,311 (6.3)** |
| *P* for trend | - | <0.001 | <0.001 | - | <0.001 | 0.337 | - | <0.001 | <0.001 |

**Abbreviation:** PA, primary aldosteronism;
\* The denominator was the number of patients with hypertension who underwent PA screening in that calendar year.

**Supplementary Table 5.** Annual Counts and Proportions of PA Screening and PA Diagnosis among Hypertensive Individuals among Potentially High-Risk Subgroups: Obstructive Sleep Apnea, Ischemic Stroke and Intracranial Hemorrhage.

| Year | Obstructive sleep apnea |  |  | Ischemic stroke |  |  | Intracranial hemorrhage |  |  |
| --- | --- | --- | --- | --- | --- | --- | --- | --- | --- |
|  | Hypertension | PA screening | Confirmed PA* | Hypertension | PA screening | Confirmed PA* | Hypertension | PA screening | Confirmed PA* |
| 2001 | 1,465 | 5 (0.34) | NR# | 44,116 | 128 (0.29) | 8 (6.3) | 9,426 | 50 (0.53) | 6 (12.0) |
| 2002 | 2,073 | 14 (0.68) | NR# | 70,265 | 213 (0.30) | 14 (6.6) | 14,222 | 71 (0.50) | 7 (9.9) |
| 2003 | 2,704 | 18 (0.67) | NR# | 94,461 | 288 (0.30) | 23 (8.0) | 18,813 | 79 (0.42) | 5 (6.3) |
| 2004 | 3,131 | 25 (0.80) | NR# | 117,261 | 391 (0.33) | 27 (6.9) | 23,603 | 113 (0.48) | 15 (13.3) |
| 2005 | 4,089 | 36 (0.88) | NR# | 138,618 | 545 (0.39) | 43 (7.9) | 28,321 | 151 (0.53) | 14 (9.3) |
| 2006 | 5,073 | 46 (0.91) | NR# | 155,099 | 531 (0.34) | 35 (6.6) | 32,306 | 158 (0.49) | 16 (10.1) |
| 2007 | 6,674 | 72 (1.08) | NR# | 169,235 | 709 (0.42) | 39 (5.5) | 35,549 | 172 (0.48) | 12 (7.0) |
| 2008 | 8,034 | 71 (0.88) | 9 (12.7) | 183,204 | 858 (0.47) | 41 (4.8) | 39,196 | 236 (0.60) | 15 (6.4) |
| 2009 | 9,184 | 111 (1.21) | 9 (8.1) | 192,746 | 1,042 (0.54) | 55 (5.3) | 42,020 | 285 (0.68) | 25 (8.8) |
| 2010 | 10,531 | 135 (1.28) | 12 (8.9) | 204,425 | 1,122 (0.55) | 61 (5.4) | 44,887 | 307 (0.68) | 28 (9.1) |
| 2011 | 11,334 | 141 (1.24) | 11 (7.8) | 216,043 | 1,152 (0.53) | 65 (5.6) | 47,812 | 308 (0.64) | 26 (8.4) |
| 2012 | 13,035 | 172 (1.32) | 14 (8.1) | 226,929 | 1,198 (0.53) | 53 (4.4) | 50,499 | 316 (0.63) | 31 (9.8) |
| 2013 | 14,058 | 159 (1.13) | 15 (9.4) | 235,511 | 1,320 (0.56) | 73 (5.5) | 53,167 | 386 (0.73) | 20 (5.2) |
| 2014 | 15,955 | 214 (1.34) | 22 (10.3) | 241,549 | 1,335 (0.55) | 61 (4.6) | 55,783 | 437 (0.78) | 36 (8.2) |
| 2015 | 17,188 | 217 (1.26) | 27 (12.4) | 247,489 | 1,572 (0.64) | 97 (6.2) | 58,150 | 458 (0.79) | 38 (8.3) |
| 2016 | 18,621 | 257 (1.38) | 22 (8.6) | 247,375 | 1,437 (0.58) | 80 (5.6) | 62,459 | 475 (0.76) | 34 (7.2) |
| 2017 | 16,243 | 285 (1.75) | 35 (12.3) | 251,455 | 1,511 (0.60) | 100 (6.6) | 74,281 | 635 (0.85) | 47 (7.4) |
| 2018 | 18,976 | 359 (1.89) | 37 (10.3) | 270,088 | 1,601 (0.59) | 106 (6.6) | 95,327 | 811 (0.85) | 64 (7.9) |
| 2019 | 22,708 | 356 (1.57) | 36 (10.1) | 270,807 | 1,631 (0.60) | 90 (5.5) | 115,786 | 908 (0.78) | 61 (6.7) |
| 2020 | 23,325 | 406 (1.74) | 41 (10.1) | 271,561 | 1,689 (0.62) | 115 (6.8) | 134,795 | 1,064 (0.79) | 99 (9.3) |
| 2021 | 23,542 | 388 (1.65) | 39 (10.1) | 274,186 | 1,812 (0.66) | 104 (5.7) | 152,448 | 1,222 (0.80) | 92 (7.5) |
| 2022 | 22,884 | 437 (1.91) | 37 (8.5) | 273,283 | 1,885 (0.69) | 102 (5.4) | 166,068 | 1,312 (0.79) | 102 (7.8) |
| **Total** | **270,827** | **3,924 (1.45)** | **386 (9.8)** | **4,395,706** | **23,970 (0.55)** | **1,392 (5.8)** | **1,354,918** | **9,954 (0.73)** | **793 (8.0)** |
| *P for trend* | - | <0.001 | 0.711 | - | <0.001 | 0.908 | - | <0.001 | 0.205 |

**Abbreviation:** PA, primary aldosteronism; NR, not reported;
\* The denominator was the number of patients with hypertension who underwent PA screening in that calendar year;

